# COMPARATIVE SEROLOGIC PROFILES OF HEPATITIS B VIRUS (HBV) BETWEEN HIV/HBV CO-INFECTED AND HBV MONO-INFECTED PATIENTS IN ILE-IFE, NIGERIA

**DOI:** 10.1101/2020.07.14.20153783

**Authors:** Oluwadamilola Gideon Osasona, Olumuyiwa Elijah Ariyo, Judith Oguzie, Testimony J.Olumade, Uwem George, Opeoluwa Adewale-Fasoro, Oluwatosin Oluwagbenga Oguntoye

## Abstract

**Introduction:** Hepatitis B virus(HBV) infects about 2 billion people globally and accounts for mortality of about 800,000 from liver cirrhosis and hepatocellular carcinoma. Sub-Saharan Africa accounts for 70% of Human Immunodeficiency Virus (HIV) global burden. HIV/HBV co-infection results in early development of HBV complications, alterations of serological biomarkers of HBV.

**Methods:** Two hundred and fifty patients with HIV/AIDS were screened for HBV and 20 (8%) were identified. Same number of HBV mono-infected individuals were recruited into the study and subsequently, HBV serological profiles which includes HBsAg, HBsAb, HBeAg, HBeAb,HBcAbIgM and HBcAbIgG were assayed using HBV ELISA kits.

**Result:** Mean age of patients in the HBV/HIV cohort was 45.5 years while the HBV mono-infected infected cohort was 30.5 years. Majority of the HBV/HIV co-infected individuals were females (85%). Frequency of HBeAg among HIV/HBV co-infected cohort was 25% and 15% for HBV mono-infected, while the frequency of HBeAb was higher (60%) among cohort of HBV/HIV co-infected patients in comparison with the HBV mono-infected cohorts(50%). Two patients among the HIV/HBV co-infected cohort have the isolated anti-HBcAg serologic pattern.

**Conclusion:** The study broadened the available evidence of comparative serologic profiles of Hepatitis B virus between cohorts of HBV/HIV co-infected individuals and HBV mono-infected patients in Nigeria.

## Introduction

There are about 2 billion people infected with hepatitis B virus (HBV) worldwide and up to 300 million are chronic carriers. This results in up to 800,000 deaths annually due to complications of liver cirrhosis and hepatocellular cancer[1,2]. Sub-Saharan Africa and Asia accounts for majority of the cases of chronic HBV infection, this has been termed areas of high endemicity with prevalence of 8% and above[3,4]. Human Immunodeficiency Virus (HIV) infection is also very common in sub-saharan Africa and accounts for more than 70% of the global burden of the disease[5], therefore, it is not uncommon to see co-infection of HIV and HBV in this area.The prevalence of HIV/HBV co-infection in sub-Saharan Africa ranges from 3.5% to 28%[8]. Nigeria has a high variation in the prevalence of HIV/HBV co-infection among people living with HIV and ranges from 10% to 70% among different subgroups of HIV positive individuals[9].This is not surprising considering similarities in the routes of transmission of HIV and HBV as both could be blood-borne and sexually transmitted[9].The wide availability of HAART also mean that more HIV patients are surviving which may contribute to the overall population of HBV co-infected patients[10].

Hepatitis B virus co-infection with HIV has been shown to worsen the outcome of HIV by accelerating progression to AIDS, HBV X-protein (HBVx) is implicated as one of the factors responsible[10,11]. Conversely, HIV could lead to early development of HBV complications[11]. Another influence of HIV on co-infection with HBV is in the alteration of biomarkers of the latter. Hepatitis B surface antigen (HBsAg) is routinely used to screen for HBV infection, while the presence of antibodies to the hepatitis B core antigen (anti-HBc) denote prior exposure to the virus. Hepatitis e antigen is a marker of infectivity and its presence correlates with high replication of the virus[12]. The presence or absence of different combination of the serological biomarkers of HBV can be used in characterising the phases of infection[13]. The phases of HBV infection could be altered by the presence of HIV and this may result in challenges with the interpretations of serology patterns. This may be due to the presence of unusual pattern in serologic profiles such as inability to detect hepatitis B core antibody or recurrence of active serology markers[14]. In some instances there were higher frequency of changes in serology biomarkers in HIV/HBV co-infected cohorts[15]. This has great implications for patient management vis-à-vis disease monitoring and follow-up. In Nigeria, some studies have assessed the serological profile of HIV/HBV co-infected patients but studies comparing this with HBV mono-infected individuals are uncommon[16,17]. This study seeks to compare the serologic profiles of HIV/HBV co-infected patients on HAART with patients who are HBV mono-infected.

## Methodology

### Study site, enrolment of participants and sample collection

This hospital-based cross-sectional study was carried out as previously described among HIV/HBV co-infected individuals and HBV mono-infected patients attending the virology clinic at the Obafemi Awolowo University Teaching Hospital, Ile-Ife, Nigeria from May to August 2019[29]. The Institutional Review Board(IRB) of the Obafemi Awolowo University Teaching Hospital, Ile-Ife granted ethical approval (reference number: NHREC/27/02/2009a) before study commenced.The study design conformed to the 2013 declaration of Helsinki. Structured questionnaire was used for the collection of demographic data, clinical information, and subsequently, blood samples were collected from each participant who consented to participate in the study.

### Laboratory Analysis

#### Detection of HBsAg, HBsAb, HBeAg, HBeAb, HBcIgG and HBcIgM antibody

A total of 250 HIV positive serum samples from patients who consented to participate in this study were initially tested for HBV using one step HBsAg strip (ACON Laboratories incorporated, USA) and 20 serum samples were found positive. Another 20 serum samples previously tested positive for HBV at the centre were also re-screened using one step HBsAg strip. All the positive samples from both HIV/HBV positive cohort, HBV mono-infected patients for one step HBsAg strip test were further tested using specific Enzyme-Linked Immunosorbent Assay (ELISA) for HBV serological markers including; HBsAg, HBsAb, HbcAb-IgG, HBcAb-IgM, HBeAg, HBeAb (MELSIN Diagnostic kits China). Both the ELISA and one step HBsAg strip assay were done according to the manufacturer’s instructions. Optical density (OD) was read using the Emax endpoint ELISA microplate reader (Molecular Devices, California, USA) and the interpretation was made in line with the manufacturer’s instructions.

### Statistical Analysis

Accuracy and completeness of questionnaires were checked, data were double entered to reduce data entry errors and later merged. All categorical data and median (interquartile range, IQR) of continuous variables were compared by Chi-square and Mann-Whitney tests, respectively, using SPSS version 25.0 (IBM, USA). The optial density values(O.D) values of HBsAg, HBsAb, HBeAg, HbeAb, HBcIgM and HBcIgG between the two study groups were also compared by Mann-Whitney test using GraphPad Prism (version 8.4.2, 2020). For all statistical analyses, P-value less than 0.05 was considered significant at 95% C.I.

The figures (Figures 1,2,3,4,5) representing the statistical significance of the serological profiles among both HIV/HBV co-infected patients and HBV mono-infected patients are presented as geometric mean ± geometric standard deviation. Only HBeAb and HBcAb are significantly different between the two study groups, according to our analysis.

**Figure 1:**
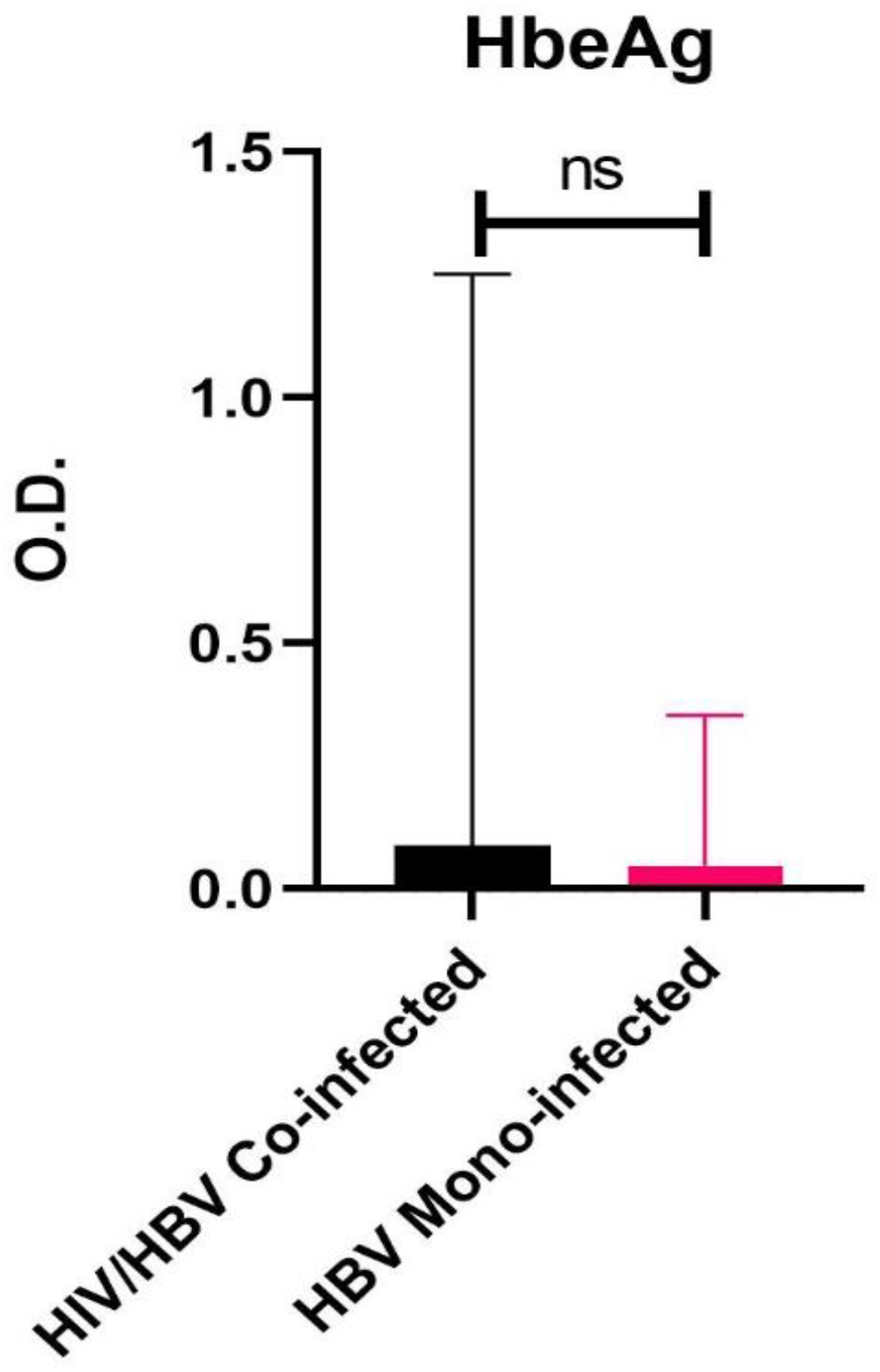
Bar plot of optical density (O.D) values of HBeAg among HIV/HBV co-infected and HBV mono-infected patients

**Figure 2:**
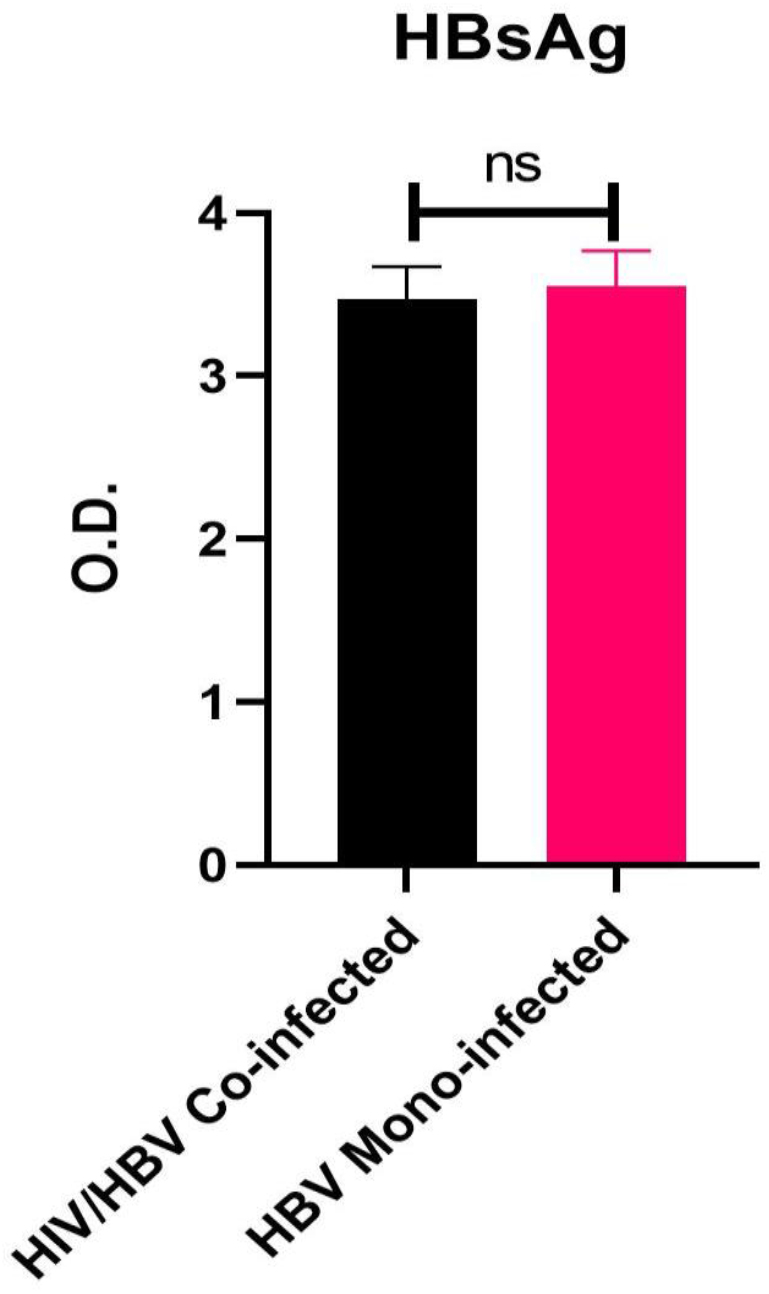
Bar plot of O.D. values of HBsAg among HIV/HBV co-infected individuals and HBV mono-infected individuals

**Figure 3:**
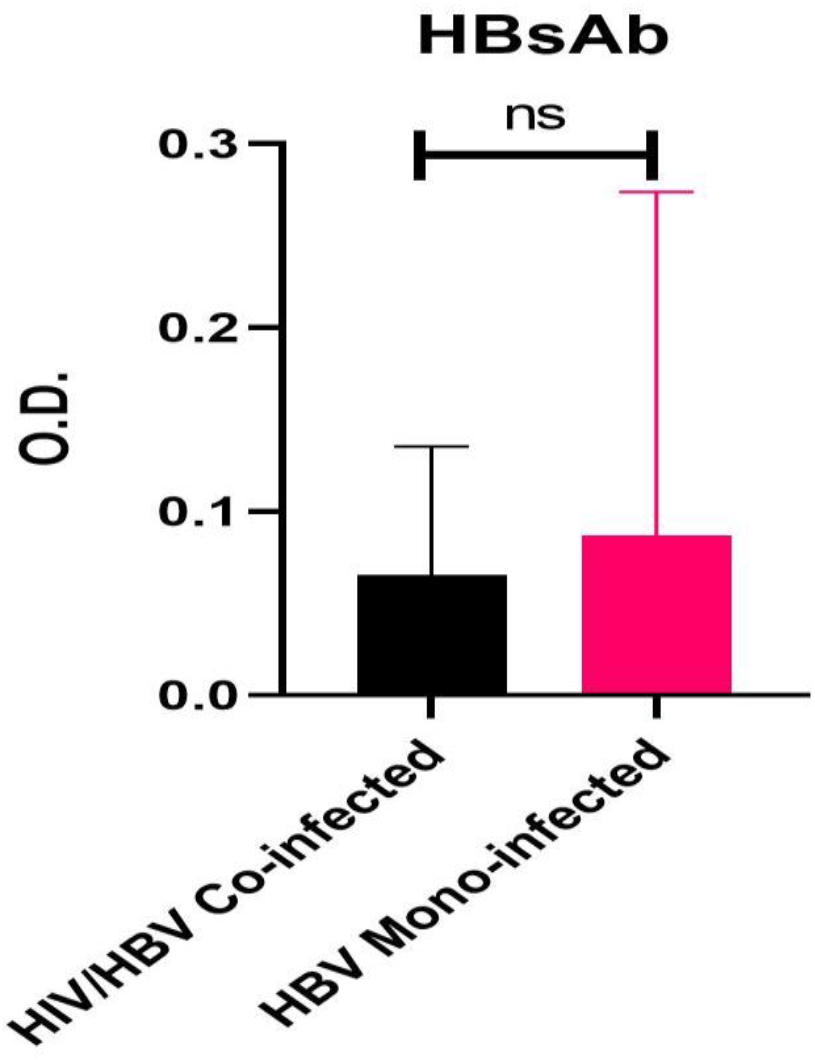
Bar plot of O.D values of HBsAg among HIV/HBV co-infected and HBV mono-infected patients

**Figure 4:**
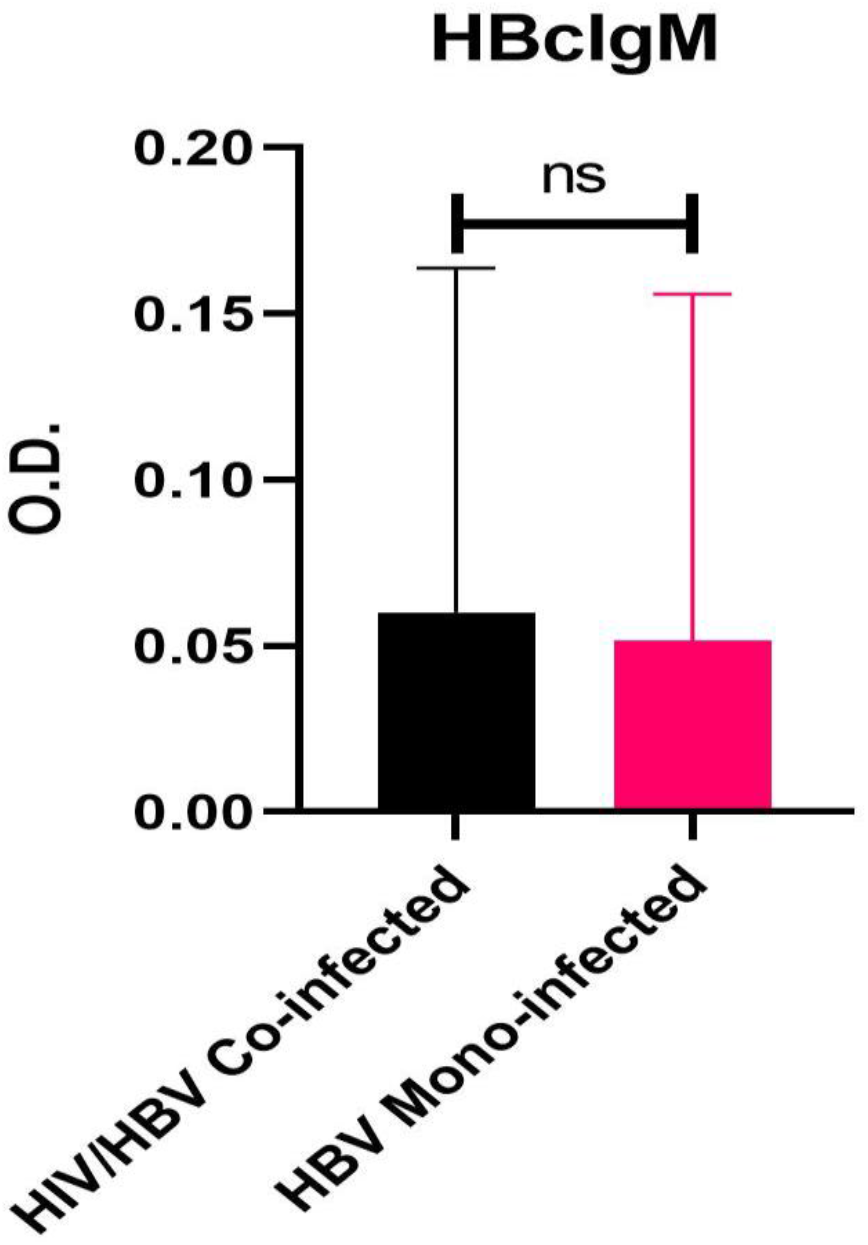
Bar plot of O.D. values of HBcIgM among HIV/HBV co-infected and HBV mono-infected individuals

**Figure 5:**
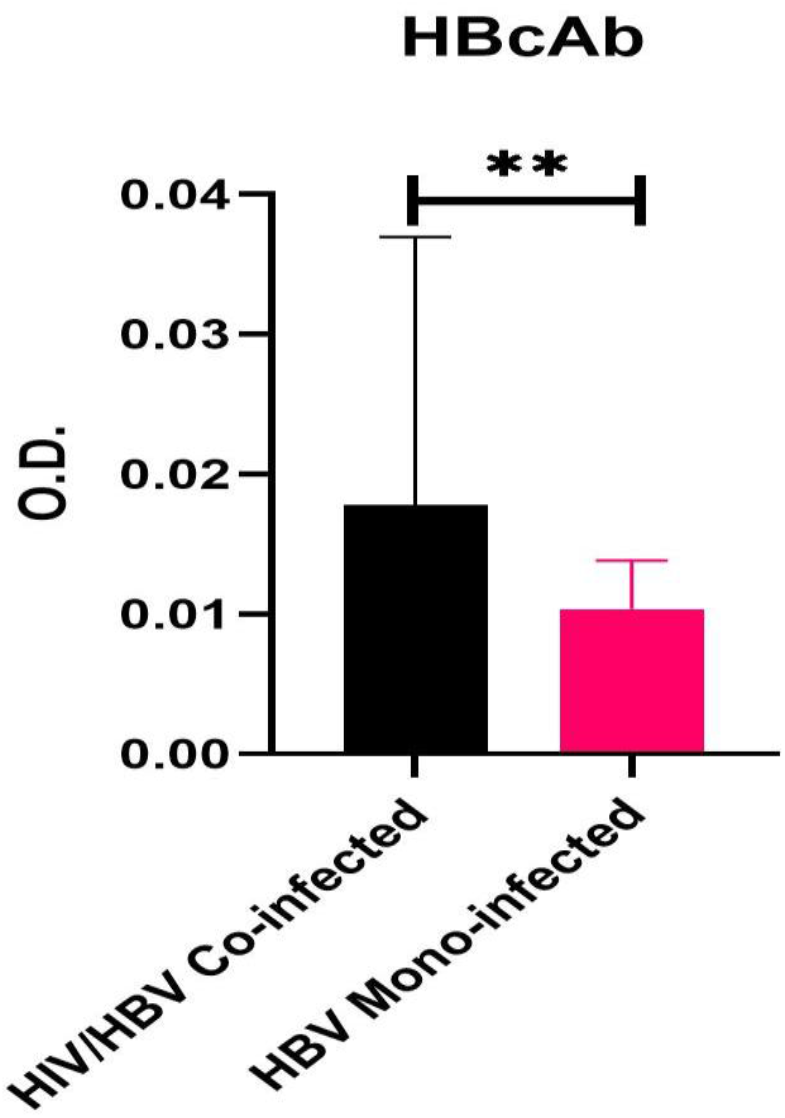
Bar plot of O.D.Values of HBcIgG among HIV/HBV co-infected and HBV mono-infected patients.

## Results

The mean age of patients in the HBV/HIV co-infected cohort was 45.5years while the median age is 40years. In the HBV mono-infected cohort, the mean age is 31.2 years and median age is 30.5 years as shown in Table 1 below. Majority of patients in the HIV/HBV co-infected cohort were females(85%) whereas more than half of the patients (60%) in the HBV mono-infected cohorts were males. Interestingly, all the 20 HIV/HBV co-infected patients in this study were unmarried or divorced unlike the HBV mono-infected cohorts where 7(35%) patients were married. Also, none of the patients in the HIV/HBV cohorts were vaccinated against HBV infection while only 2 (10%) of the HBV mono-infected were vaccinated against HBV infection. A large proportion (85%) of HIV/HBV co-infected patients had prior history of circumcision (both male and female) while 60% of HBV mono-infected patients had been circumcised using unsterilized objects in the past as depicted in the table 1 below,however, this is not statistically significant (p value = 0.077) Two hundred and fifty patients living with HIV were screened for HBsAg using HBV rapid diagnostic kit and HBV ELISA test kits. Twenty (8%) tested positive. Same number of HBV mono-infected patients (20) were recruited into the study as controls after testing using HBV ELISA kits.

**Table 1:**
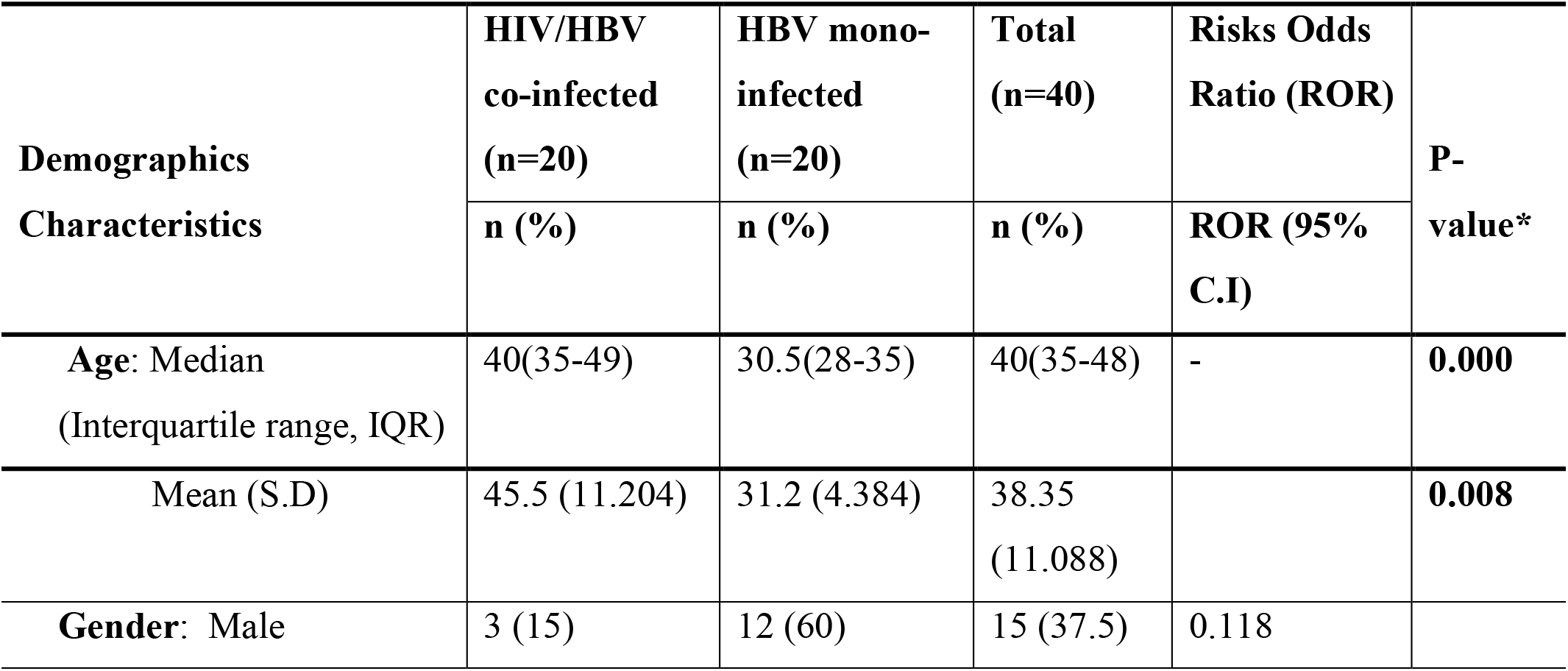
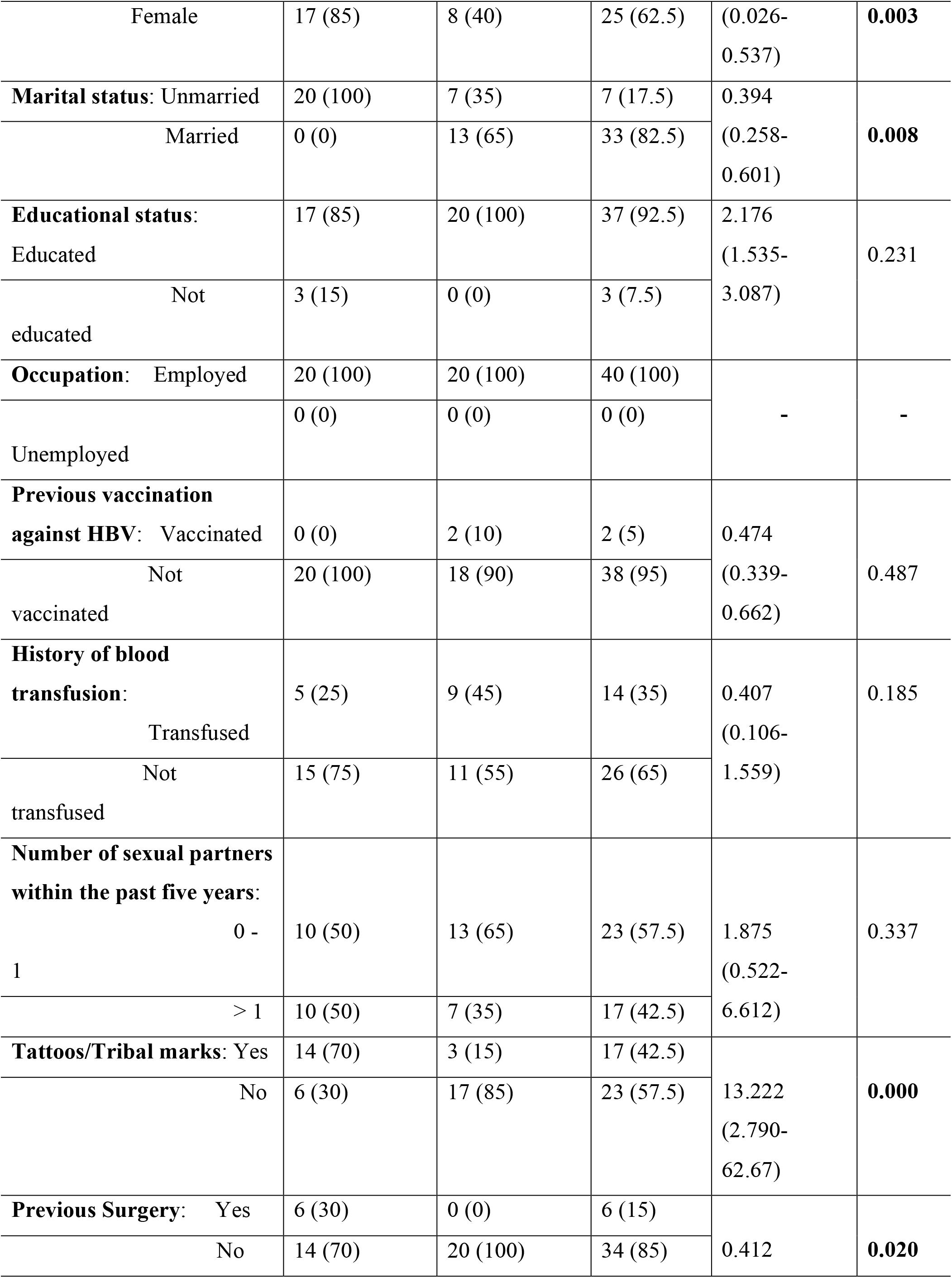
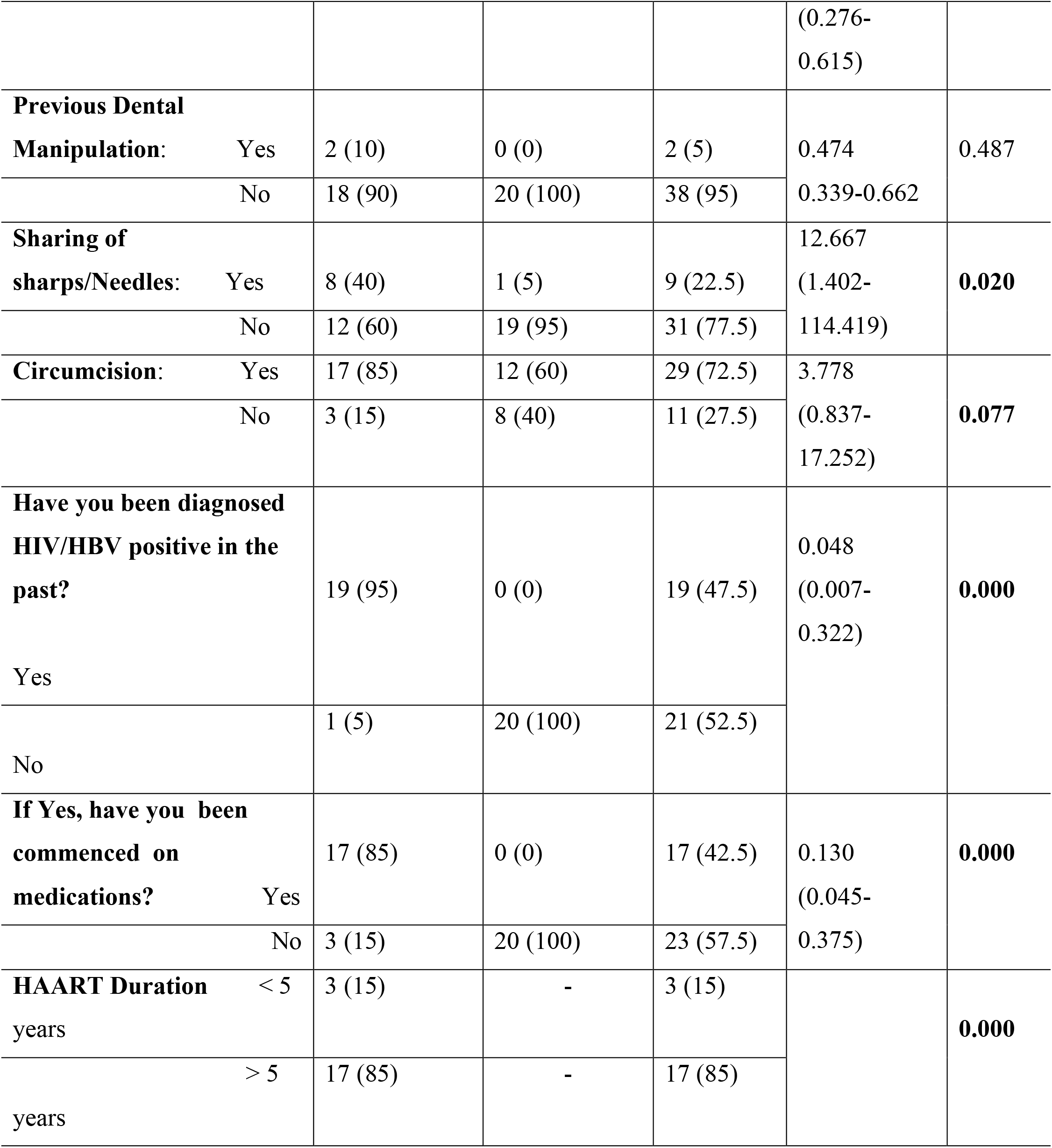
Demographic characteristics of study subjects

The hepatitis B virus serological profiles of all the patients in each cohort were assayed using HBV ELISA kit for HBsAg, HBsAb, HBeAg, HBeAb, HBcIgM, HBcIgG. Five (25%) of the HBV/HIV co-infected patients and 3 (15%) of the HBV mono-infected were positive for HBeAg(Figure 1 and 6), Fifty percent of the HIV/HBV co-infected were positive for HBeAb while 60% were positive in the HBV mono-infected cohort(Figure 6). HBcIGM assay of the HIV/HBV co-infected cohort yielded 15% while the mono-infected yielded 10% (Figures 4 and 6). Of the HIV/HBV co-infected, all (100%) were positive for HBcIgG Ab while 19 (95%) were positive in the HBV mono-infected cohort (Figures 5 and 6). Two (ten percent) of the HIV/HBV co-infected patients were postive for HBsAb while 4 (20%) were positive in the HBV mono-infected cohort(Figures 3 and 6). Majority of patients (85%) with HIV/HBV co-infection had been on highly active antiretroviral therapy for more than 5 years. We also observed the presence of isolated anti-HBcAb in two (10%) patients among the HIV/HBV cohort of patients. This particular serologic finding was not observed among the HBV mono-infected cohort.

**Figure 6:**
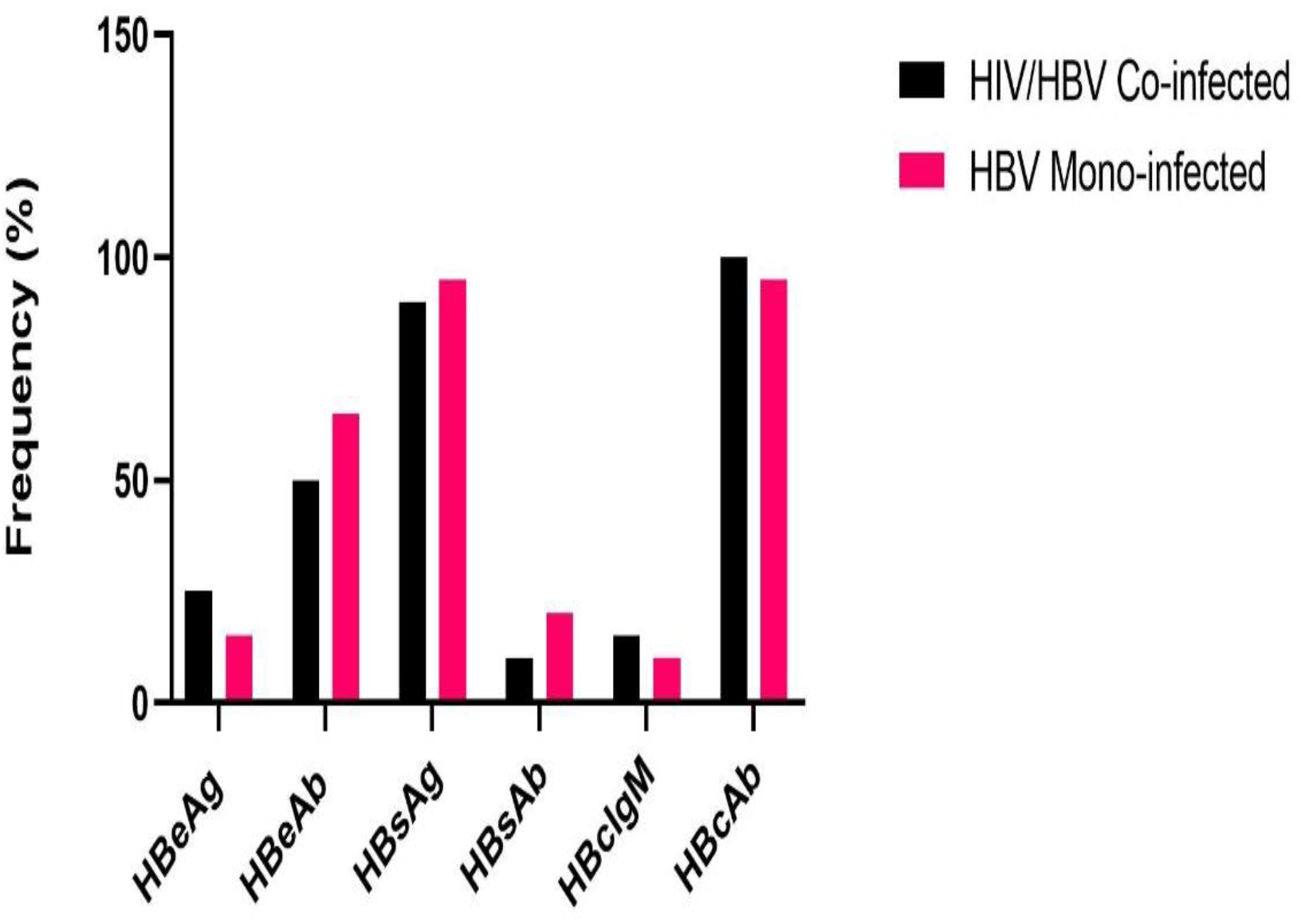
Serological profiles of HBV between HIV/HBV co-infected and HBV mono-infected patients.

## Discussion

The prevalence of HBsAg among HIV/HBV co-infected patients seen at the clinic was 8% and the patients were observed to be older in comparison to the HBV mono-infected cohort with mean ages 45.5years and 31.2 years respectively. This might be explained by the fact that in hyperendemic region for HBV infection such as Asia and sub-Saharan Africa, patients get infected at relatively younger ages mostly at childhood[18]. This is not unexpected due to high rate of perinatal and horizontal transmission of HBV in hyperendemic countries(>8%)[23], high infectivity rate of HBV[18], associated risk factors commoner in younger populations.Most of the patients with HBV/HIV co-infection were females (85%). This may be due to gender differences in health seeking behavior among different ethnic groups in different regions of the country which is attributable to socio-cultural beliefs and norms in various regions[19].The observed prevalence of HBsAg among co-infected patients in this study falls below the national prevalence of HBsAg among people living with HIV/AIDS which ranges from 16-52%[20] and higher than the West African regional prevalence of 6.1%[21] and global prevalence of 7.4%[22].

High rate of circumcision (85% and 60% respectively) especially female circumcision was observed from the study. This practice is usually carried out by women in traditional Yoruba settings using instruments whose sterility cannot be guaranteed thereby increasing the risk of contacting blood borne viruses like HIV and HBV. Although, there has been an increase in national campaign in Nigeria to stop this harmful female traditional practices but it is still being carried out in hinterlands in some regions in the country[24].

The prevalence of HBeAg among HIV/HBV co-infected patient in the study was 25% and higher than that of HBV mono-infected patients (15%) as shown in figures 1 and 6 above, although not statistically significant. The observed prevalence of HBeAg among HIV patients from this study is comparatively higher than the findings by Awoderu et al[26] in Lagos, where they observed a prevalence of 3.1% among HIV positive patients.This high prevalence of HBeAg is not unexpected as HIV/AIDS infection confers a state of immunosupresion on the host with resultant increased rate of replication of the HBV. Also, there is documented evidence of seroreversion of HBeAb to HBeAg among patients with HBV/HIV co-infection which could have accounted for the increased level of HBeAg[25]. Of importance also, is the fact that the HIV/HBV cohort have higher frequency of HBcIgM in comparison with the HBV mono-infected which is depicted in figures 4 and 6 above, however the relationship is not statistically significant. This confirms that acute HBV infection (< 6 months) is higher in this cohort with possibility of progression to a chronic state as evidenced by higher frequency of HBcIgG antibody among cohort of HBV/HIV co-infected patients (figures 5 and 6). We also observed the “isolated anti-HBcAb” in two (10%) of the patients with HIV.This set of patients have higher prevalence of occult hepatitis B infection. Opaleye et al in Nigeria observed a prevalence of 29.2% of isolated anti-HBcAb among HIV patients[28] and from this cohort of patients with isolated anti-HBcAb, 28.6% progressed to develop occult hepatitis B virus infection[28].Piroth et al in a longitudinal study on evolution of hepatitis B virus serological profiles among HIV positive patients in France over 36 months observed persistent isolated anti-HBc reactivity in 35% of the patients[27].This finding may have implication for the correct interpretation of serologic results among HIV/HBV co-infected patients and therefore necessitate the isolation of HBV DNA in this cohort of patients for proper diagnosis.This study also established an understandably comparative low prevalence of HBsAb among HIV/HBV co-infected individuals in comparison to the HBV mono-infected cohort(10% vs 15%).This pattern is similar to findings by Toscano et al in Brazil[25]. The reasons advanced for this findings above include quantitative and qualitative defect in HBsAb production due to immunosuppression by HIV, seroreversion of HBsAb to HBsAg[25].

The limitation of this study includes inadequate sample size which may affect the power of the study and therefore the inference made to the wider population, it is recommended that more patients should be enrolled into future studies. Also, inclusion of the CD4 count, liver enzymes, antiretroviral therapy(ART) status of patients and viral load would enable researchers to make a robust inference from the observed serological profiles of HBV between the two cohorts of patients studied.

## Conclusion

The study has enhanced the available repertoire of comparative serologic profiles of hepatitis B virus among cohorts of HIV/HBV co-infected individuals in comparison to HBV mono-infected cohorts in Nigeria and globally. It further confirmed the higher frequency of HbeAg, HBcIgM, HBcIgG and lower frequency of HBeAb, HBsAb among cohorts of HIV/HBV co-infected patients in comparison to HBV mono-infected individuals. We observed the presence of isolated anti-HBcAb among HIV positive patients. We therefore recommend that complete panel of HBV serological profiles be routinely assayed in patients with HBV/HIV co-infection in Nigeria.

## Data Availability

The data for this study are available from the corresponding author
upon request

## Author’s Contributions

Conceptualisation/Design of research study – O.G.O., U.M., O.A.F Data curation : O.G.O., T.J.O

Formal Analysis : T.J.O, J.O. Resources : O.G.O., O.A.F.

Writing Original Draft : O.G.O., U.G.,O.E.A,O.A.F Writing Review and Editing : O.O.O., J.U., O.G.O.

All authors have read and approved the final manuscript.

## Author information

O.O.O – Senior Lecturer/Consultant Gastroenterologist(FWCP)

O.E.A – Senior Registrar, Internal Medicine/Infectious Diseases(MSc Immunology,United Kingdom)

O.G.O – Supernumerary Resident, Internal Medicine/MSc Virology(Ibadan, Nigeria), MSc Infectious Diseases(LSHTM,United Kingdom),Ph.d Infectious Diseases Genomics(in view, ACEGID, Nigeria)

U.G – MSc Virology(Ibadan, Nigeria), Ph.d Infectious Diseases Genomics(in view, ACEGID, Nigeria)

T.J.O – MSc Molecular Biology and Genomics (Nigeria), Ph.d Infectious Diseases Genomics(in view, ACEGID, Nigeria)

O.A.F-MSc student at department of Molecular Microbiology and Immunology at the Johns Hopkins Bloomber School of Public Health.

## Acknowledgements

We are deeply grateful to Dr MO Adewumi and Dr K Adeniji of the Department of Virology, University of Ibadan, Nigeria for provision of laboratory resources and their excellent technical consultation.

